# Uptake and safety of pneumococcal vaccination in adults with immune mediated inflammatory diseases: a nationwide observational study using data from the Clinical Practice Research Datalink (Gold) in the UK

**DOI:** 10.1101/2023.12.13.23299925

**Authors:** Georgina Nakafero, Matthew J. Grainge, Tim Card, D. Mallen Christian, Jonathan S. Nguyen Van-Tam, Abhishek Abhishek

## Abstract

**Introduction:** The uptake and safety of pneumococcal vaccination in people with immune mediated inflammatory diseases (IMIDs) is poorly understood. We investigated the UK wide pneumococcal vaccine uptake in adults with IMIDs and explored the association between vaccination and IMID flare.

**Methods:** Adults with IMIDs diagnosed on or before 01/09/2018, prescribed steroid-sparing drugs within the last 12 months and contributing data to the Clinical Practice Research Datalink Gold were included. Vaccine uptake was assessed using a cross-sectional study design. Self-controlled case series (SCCS) analysis investigated the association between pneumococcal vaccination and IMID flare. The SCCS observation period was up-to six-month before and after pneumococcal vaccination. This was partitioned into 14-day pre-vaccination, 90-days post-vaccination exposed, and the remaining unexposed period.

**Results:** We included 32,277 patients, 14,151 with RA, 13,631 with IBD, 3,804 with axial spondyloarthritis and 691 with SLE. Overall, 50% of patients were vaccinated against pneumococcus. Vaccine uptake was lower in those younger than 45 years (30%), with IBD (38%), and without additional indication(s) for vaccination (43%). In the vaccine-safety study, data for 1001, 854, 424 vaccinated patients with primary-care consultations for joint pain, AIRD flare and IBD flare respectively were included. Vaccination against pneumococcal pneumonia was not associated with primary-care consultations for joint pain, AIRD flare and IBD flare in the exposed period with incidence rate ratios (95% Confidence Interval) 0.95 (0.83-1.10), 1.07 (0.93-1.22), and 0.82 (0.64-1.06) respectively.

**Conclusion:** Uptake of pneumococcal vaccination in UK patients with IMIDs was suboptimal. Vaccination against pneumococcal disease was not associated with disease flare.

## Introduction

Immunosuppressed adults with immune-mediated inflammatory diseases (IMIDs) such as rheumatoid arthritis (RA), inflammatory bowel disease (IBD), and systemic lupus erythematosus (SLE) are at an increased risk of pneumonia, and its complications including death^1–4^. Consequently, pneumococcal vaccination is recommended for this at-risk population^5 6^. Despite long-standing recommendations for vaccinating the risk groups since 1992 and the over 65s since 2003^5^, the uptake of pneumococcal vaccination in the at-risk population is suboptimal^7^. In a previous study from the UK, the uptake of pneumococcal vaccination among patients with RA was reported to be 50% overall, and 43% in those younger than 65 years in age^8^. The uptake of pneumococcal vaccination across a broad range of IMIDs in a UK wide cohort has not been evaluated to the best of our knowledge. Understanding vaccine uptake across a range of conditions is important as the uptake of pneumococcal vaccination in people with IBD was noted to be lower in North America and Europe at 10.3%-38% ^9 10^.

Belief that the vaccination could trigger an IMID flare, and cause other IMIDs e.g., vasculitis are key barriers to vaccination^9 11 12^. The association between pneumococcal vaccination and IMID flare has not been evaluated in an adequately controlled study. There is some evidence from small uncontrolled studies restricted to a few conditions that vaccination against pneumococcal disease does not cause a flare of IMIDs. In a systematic review and meta-analysis of pneumococcal vaccine immunogenicity studies in patients with SLE, disease activity did not worsen up to eight weeks after pneumococcal vaccination^13^.

In this study we evaluated the uptake and safety of pneumococcal vaccination in adults with IMIDs.

## Methods

### Data source

Data from the Clinical Practice Research Datalink (CPRD) Gold were used in this study. Incepted in the year 1987, CPRD Gold is an anonymised longitudinal database of electronic health records of >14 million people in the UK. CPRD participants are representative of the UK population in terms of age, sex, and ethnicity^14^. CPRD includes information on demographics, lifestyle factors, diagnoses stored as Read codes – a coded thesaurus of clinical terms, primary-care prescriptions, and immunisations. Vaccination and date of vaccination are also recorded.

### Approval

CPRD Research Data governance (Reference 21_000614).

### Study design

Cross-sectional and self-controlled case series (SCCS) study designs were used to examine the uptake and safety of pneumococcal vaccination.

### Population

Adults aged ≥18 years on the 1^st^ September 2018, with at-least one primary-care record of an IMID (i.e., rheumatoid arthritis (RA), inflammatory bowel disease (IBD), axial spondyloarthritis (Ax-SpA), systemic lupus erythematosus (SLE)) and with at least one prescription of a steroid sparing drug (i.e., either methotrexate, azathioprine, 5-mercaptopurine, sulfasalazine, 5-aminosalicylates, mycophenolate, leflunomide, ciclosporin, tacrolimus or sirolimus) within the previous 12 months were eligible to be included in this study.

### Pneumococcal vaccination

Pneumococcal vaccination was defined using both product codes and Read codes (Table S1). Dates of vaccination were extracted from CPRD. Data on vaccination from inception of CPRD in 1987 to 1^st^ September 2018 were considered in the vaccine uptake study. Vaccinations recorded in the CPRD as not administered in primary care e.g., vaccination from hospitals, pharmacy were included in the vaccine uptake study but were excluded from the safety study because the date of administration is not reliably recorded in the CPRD. Similarly, as the UK vaccination policy is to offer a single dose of vaccination against pneumococcal pneumonia, we only considered the latest vaccination record in those with two or more records of vaccination.

### Outcomes

#### Vaccine uptake

Vaccination against pneumococcal pneumonia.

#### Vaccine safety

A. Auto-immune rheumatic disease (AIRD)

[1] Primary care consultation for joint pain. This was defined using Read codes (Table S2). Consultations for joint pain within 14 days of each other were considered as part of the same episode.
[2] AIRD flare. This was defined as present when there was a primary-care prescription of oral corticosteroid without another corticosteroid prescription in the preceding sixty days. The patient was also required to not have consulted for an alternate condition that could justify corticosteroid prescription on the same date. For this, all relevant primary care consultations were retrieved and reviewed by AA (General Medicine and Rheumatology expertise) for conditions that might explain the corticosteroid prescribed and such participants were excluded from the analysis as there was considerable uncertainty whether they experienced IMID flare or another illness (Table S3). The corticosteroid prescription free period used to define consultation for AIRD flare was increased to at-least 120 days in a sensitivity analysis as this time-period has been validated for the IBD flare^15^ (See below).
B. IBD

[1] IBD flare. This was defined as present when there was primary-care prescription of corticosteroid without another corticosteroid prescription in the preceding 120 days^15^. The patient was also required to not have consulted for an alternate condition that could justify corticosteroid prescription on the same date defined as a new primary care prescription of corticosteroid^15^.

### Covariates

#### Vaccine uptake

Age, sex, type of IMID and presence of additional indication for vaccination^5^. Briefly, these included chronic heart diseases, chronic respiratory diseases, chronic kidney diseases, chronic liver diseases, immunosuppression, diabetes and asplenia.

#### Vaccine safety

Season defined in line with the Meteorological Office. According to the Meteorological Office, winter spans from the 1^st^ December to the 28^th^ February of the next year, spring spans from 1^st^ March to 31^st^ May, summer spans from 1^st^ June to 31^st^ August, and autumn spans from 1^st^ September to 31^st^ November.

### Statistical analyses

#### Vaccine uptake

The percentage and 95% CI of the study population alive on 1^st^ September 2018 that were vaccinated was calculated. The proportion vaccinated was stratified according to their age (<45, 45–64, ≥65 years) on the 1^st^ of September 2018, sex, presence of other indications for vaccination, and type of IMID (RA, IBD, SLE, Ax-SpA). Logistic regression was used to examine mutually adjusted associations between pneumococcal vaccination and age group, sex, IMID type and presence of additional at-risk condition for vaccination.

#### Vaccine safety

Patients vaccinated against pneumococcal pneumonia and who also consulted their GP for at-least one IMID flare in the six-month period before and the six-month period after vaccination were included in a self-controlled case-series (SCCS) analysis. SCCS is an established study design for assessing vaccine safety. By including patients with both an exposure and an outcome, and undertaking within person comparisons, it removes between person confounding, a key confounder in vaccine safety studies. The baseline period extended from the latest of current registration date, first IMID diagnosis date recorded in the CPRD, and six-months preceding vaccination to 15-days pre-vaccination, and from 90 days post-vaccination to the earliest date of six-months post vaccination, leaving GP surgery, death, or last data collection from the GP surgery. The exposed period extended from vaccination date to 90 days later and, was further categorised as 0-14 days, 15-30 days, 31-60 days, and 61-90 days. The first cut-off was selected at 14-days post-vaccination as it takes two weeks for the serological response and, we hypothesised this period of immune reconstitution would be most likely to associate with disease activity. The 15-day period immediately preceding vaccination was excluded from the baseline period to minimise confounding due to healthy vaccinee effect or due to active promotion of vaccination in those consulting for a disease flare^16^.

A Poisson model conditioned on the number of events adjusted for the four UK seasons as categories was fitted to calculate incidence rate ratios (aIRR) and 95% confidence interval (CI) for each exposure period compared to the baseline period. Data management and analysis were performed in Stata v17, Stata Corp LLC, Texas, USA.

## Results

### Uptake

Data from 32,277 people with IMIDs were included in this study (Figure 1). Their mean age (SD) on 1^st^ of September 2018 was 58 (16) years and 59% were female. 14,151 (43.8%) had RA, 13,631 (42.2%) IBD, 3,804 (11.8%) Ax-SpA, and 691 (2.1%) SLE.

**Figure 1:**
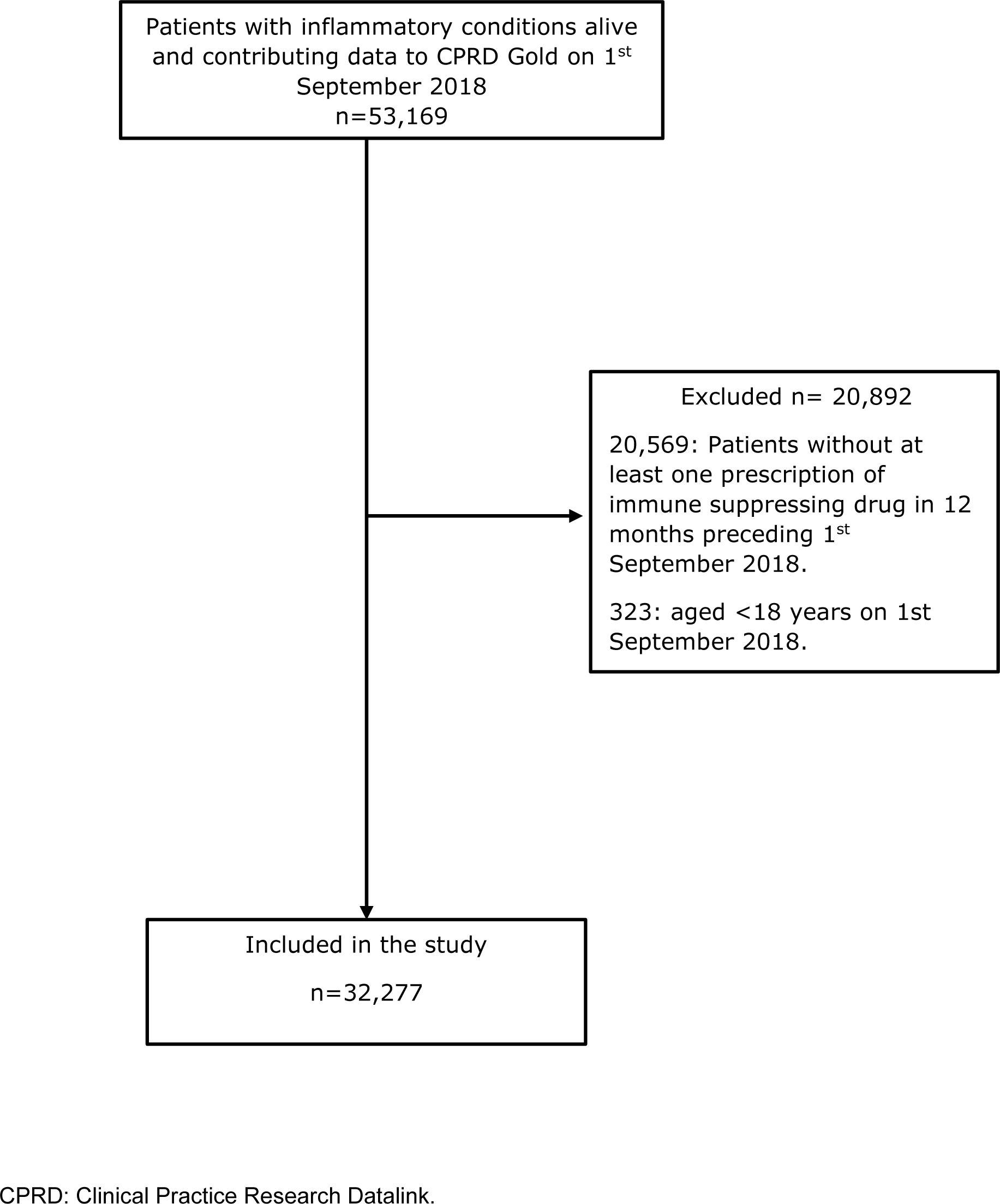
Participant selection criteria.

Overall uptake of pneumococcal vaccination was 50% (95%CI 49.4-50.5%). Vaccination uptake was significantly lower in people with IBD (38.4%, 95% CI (37.5-39.2%)) than in AIRDs (58.5%, 95% CI (57.8-59.2%)) with adjusted odds ratio (aOR) 0.57 (95%CI 0.54-0.59)) (Table 1). Patients with IBD and SLE were significantly less likely to be vaccinated compared to those with RA.

**Table 1:**
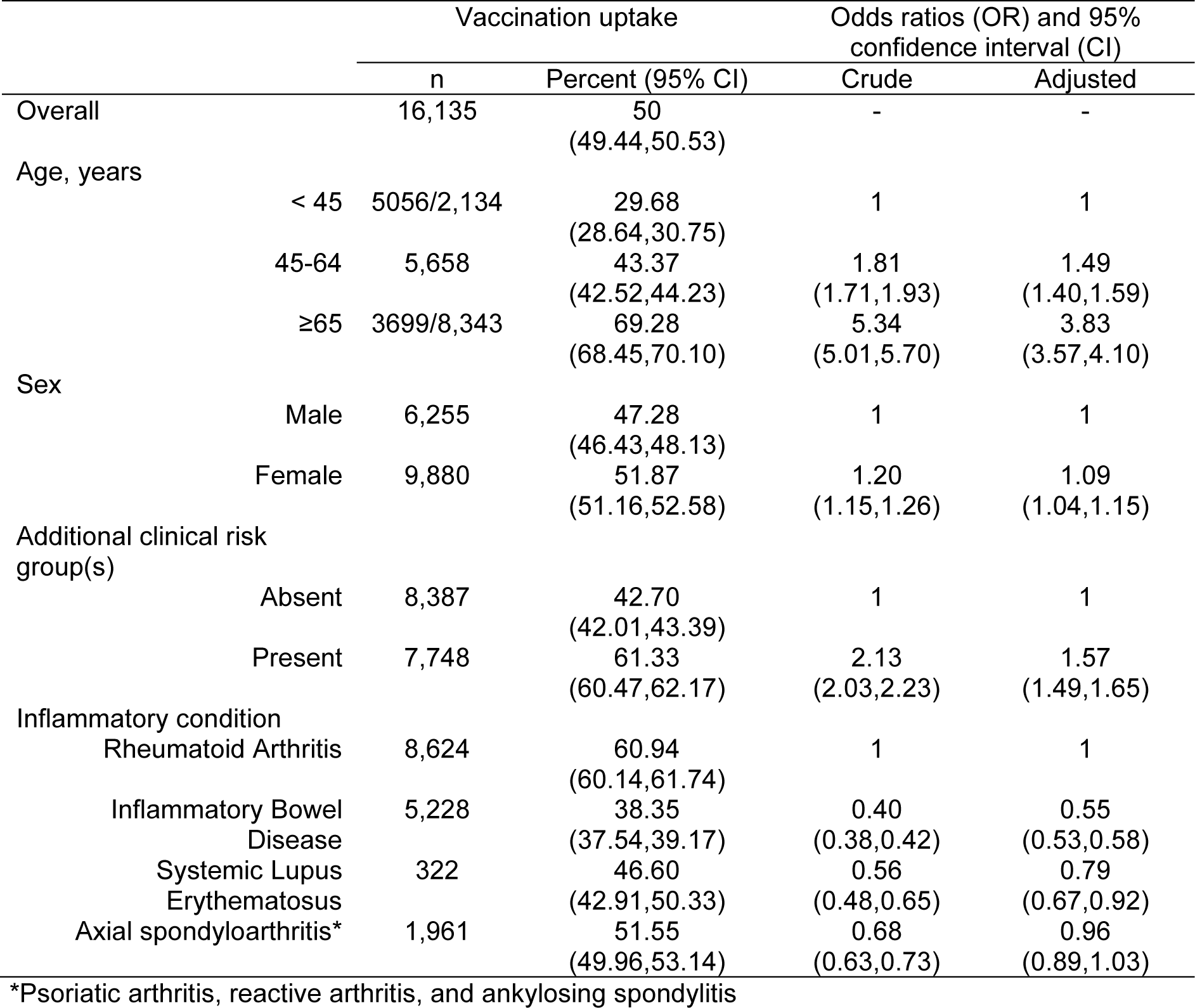
Uptake and risk factors of pneumococcal vaccination in immune mediated inflammatory diseases (n=32,277)

Increasing age, female sex and presence of additional at-risk conditions were independently associated with the uptake of pneumococcal vaccination. Vaccination uptake was higher in people aged at-least 65 years or with an additional at-risk condition (uptake (95% CI) 62.7(62.0-63.4)) than in those less than 65-years in age and without an additional at-risk condition for vaccination (uptake (95% CI) 34.2 (33.1-35.1)). On adjusting for gender and type of inflammatory condition, people not considered at additional risk of pneumococcal pneumonia (i.e., age < 65 years and/or without another at-risk condition) were 68% less likely to get vaccinated than those considered at additional risk of pneumococcal pneumonia (i.e., aged ≥65-year and/or with additional at-risk condition) with aOR 0.32 (95%CI 0.31-0.33)).

### Vaccine Safety

Data for 1001, 854, and 424 participants with primary care consultations for joint pain, AIRD flare, and IBD flare respectively were included. 1,702 participants had either AIRD flare or a primary care consultation for joint pain and of these, 1,308 (76.9%) had RA, 266(10%) had SpA and 128 (7.5%) had SLE. The majority were female (71%) and their mean (SD) age was 55 (12) years. Of the participants with IBD flare, 224 (52.8%) had ulcerative colitis, 150 (11.8%) had Crohn’s disease, 50 (11.8%) had IBD without any specific coding for subtype.

Vaccination against pneumococcal pneumonia was not associated with joint pain consultation, AIRD flare or IBD flares respectively in the 90-days post vaccination (Tables 2, 3). The 15-day pre-vaccination period associated with significantly more primary care consultations for joint pain, AIRD flare and IBD flare (Tables 2, 3).

**Table 2:**
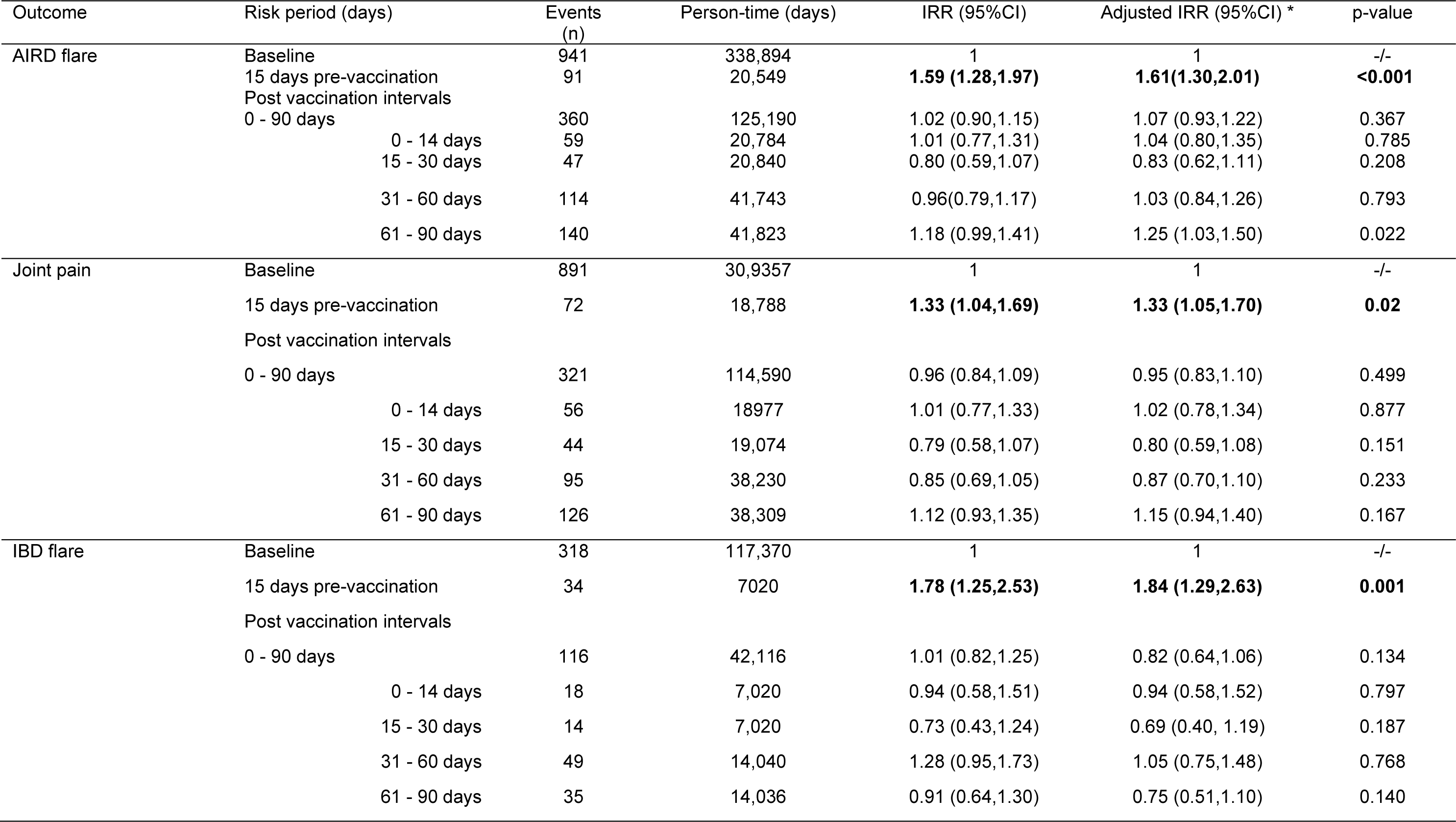
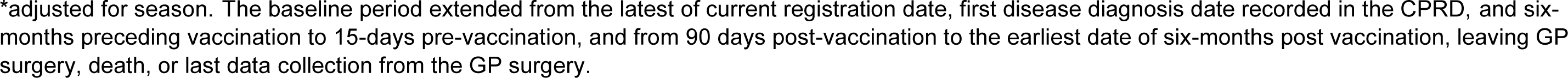
The association between pneumococcal vaccination and consultation for AIRD flare, joint pain, and IBD flare.

| Outcome | Risk period (days) | Events (n) | Person-time (days) | IRR (95%CI) | Adjusted IRR (95%CI) * | p-value |
| --- | --- | --- | --- | --- | --- | --- |
| AIRD flare | Baseline | 941 | 338,894 | 1 | 1 | -/- |
|  | 15 days pre-vaccination | 91 | 20,549 | **1.59 (1.28,1.97)** | **1.61(1.30,2.01)** | **<0.001** |
|  | Post vaccination intervals |  |  |  |  |  |
|  | 0 - 90 days | 360 | 125,190 | 1.02 (0.90,1.15) | 1.07 (0.93,1.22) | 0.367 |
|  | 0 - 14 days | 59 | 20,784 | 1.01 (0.77,1.31) | 1.04 (0.80,1.35) | 0.785 |
|  | 15 - 30 days | 47 | 20,840 | 0.80 (0.59,1.07) | 0.83 (0.62,1.11) | 0.208 |
|  | 31 - 60 days | 114 | 41,743 | 0.96(0.79,1.17) | 1.03 (0.84,1.26) | 0.793 |
|  | 61 - 90 days | 140 | 41,823 | 1.18 (0.99,1.41) | 1.25 (1.03,1.50) | 0.022 |
| Joint pain | Baseline | 891 | 30,9357 | 1 | 1 | -/- |
|  | 15 days pre-vaccination | 72 | 18,788 | **1.33 (1.04,1.69)** | **1.33 (1.05,1.70)** | **0.02** |
|  | Post vaccination intervals |  |  |  |  |  |
|  | 0 - 90 days | 321 | 114,590 | 0.96 (0.84,1.09) | 0.95 (0.83,1.10) | 0.499 |
|  | 0 - 14 days | 56 | 18977 | 1.01 (0.77,1.33) | 1.02 (0.78,1.34) | 0.877 |
|  | 15 - 30 days | 44 | 19,074 | 0.79 (0.58,1.07) | 0.80 (0.59,1.08) | 0.151 |
|  | 31 - 60 days | 95 | 38,230 | 0.85 (0.69,1.05) | 0.87 (0.70,1.10) | 0.233 |
|  | 61 - 90 days | 126 | 38,309 | 1.12 (0.93,1.35) | 1.15 (0.94,1.40) | 0.167 |
| IBD flare | Baseline | 318 | 117,370 | 1 | 1 | -/- |
|  | 15 days pre-vaccination | 34 | 7020 | **1.78 (1.25,2.53)** | **1.84 (1.29,2.63)** | **0.001** |
|  | Post vaccination intervals |  |  |  |  |  |
|  | 0 - 90 days | 116 | 42,116 | 1.01 (0.82,1.25) | 0.82 (0.64,1.06) | 0.134 |
|  | 0 - 14 days | 18 | 7,020 | 0.94 (0.58,1.51) | 0.94 (0.58,1.52) | 0.797 |
|  | 15 - 30 days | 14 | 7,020 | 0.73 (0.43,1.24) | 0.69 (0.40, 1.19) | 0.187 |
|  | 31 - 60 days | 49 | 14,040 | 1.28 (0.95,1.73) | 1.05 (0.75,1.48) | 0.768 |
|  | 61 - 90 days | 35 | 14,036 | 0.91 (0.64,1.30) | 0.75 (0.51,1.10) | 0.140 |

\*adjusted for season. The baseline period extended from the latest of current registration date, first disease diagnosis date recorded in the CPRD, and six-months preceding vaccination to 15-days pre-vaccination, and from 90 days post-vaccination to the earliest date of six-months post vaccination, leaving GP surgery, death, or last data collection from the GP surgery.

**Table 3.** The association between pneumococcal vaccination and AIRD flare^†^: Sensitivity analysis.

| Risk period (days) | Events<br>(n) | Person-time<br>(days) | IRR (95%CI) | Adjusted IRR<br>(95%CI) * | p-value |
| --- | --- | --- | --- | --- | --- |
| Baseline | 213 | 81,274 | 1 | 1 | -/- |
| 15 days pre-vaccination | 23 | 4,874 | 1.80 (1.17,2.76) | 1.99 (1.29,3.09) | 0.002 |
| Post vaccination intervals |  |  |  |  |  |
| 0 - 90 days | 96 | 29,837 | 1.20 (0.94,1.53) | 1.16 (0.88,1.53) | 0.283 |
| 0 - 14 days | 16 | 4,948 | 1.21 (0.73,2.02) | 1.32 (0.79,2.20) | 0.289 |
| 15 - 30 days | 12 | 4,969 | 0.90 (0.50,1.62) | 0.96 (0.53,1.72) | 0.887 |
| 31 - 60 days | 31 | 9,960 | 1.16 (0.80,1.69) | 1.16 (0.78,1.72) | 0.470 |
| 61 - 90 days | 37 | 9,960 | 1.39 (0.98,1.97) | 1.36 (0.94,1.97) | 0.104 |

\*adjusted for season; <sup>†</sup>AIRD flare defined as a ≥4-month gap between steroid prescriptions in people with autoimmune rheumatic diseases (AIRDs) during the observation period. The baseline period extended from the latest of current registration date, first disease diagnosis date recorded in the CPRD, and six-months preceding vaccination to 15-days pre-vaccination, and from 90 days post-vaccination to the earliest date of six-months post vaccination, leaving GP surgery, death, or last data collection from the GP surgery.

## Discussion

This UK wide study has shown that only one in two immunosuppressed adults with IMIDs in the UK is vaccinated against pneumococcal pneumonia. This is similar to the vaccine uptake of 54% to 56% reported in people with chronic respiratory disease, chronic kidney disease, and diabetes requiring insulin or oral hypoglycaemic medication. The vaccine uptake was even lower at 30% in those less than 45-years in age and at 43% in IMID patients without an additional indication for vaccination. The vaccine uptake ranged from 38.4% to 60.9% in IBD and RA respectively. This study did not find an association between pneumococcal vaccination and IMID flare requiring medical attention. An increased risk in flare of underlying disease was observed 15-days pre-vaccination, which could be attributed to opportunistic vaccine promotion to people consulting for a flare.

It is difficult to compare our findings on vaccine uptake in IMIDs with those of previous studies, since, to our knowledge, this is the first study to assess pneumococcal vaccination uptake across many inflammatory conditions and to compare uptake between different inflammatory conditions. These low vaccination rates are concerning given the increased risk of pneumococcal disease in this at-risk population for whom vaccination is recommended and indicate that they would benefit from targeted measures to increase pneumococcal vaccine uptake. We observed substantial variation in vaccination rates across inflammatory disease type. This may reflect the fact that recommendation of immunosuppression in inflammatory condition meriting vaccination is left to individual doctors’ clinical judgement, which may result in differential advice. A previous UK primary care study reported similar 50% pneumococcal vaccine uptake in RA patients^8^. The lack of difference in vaccination rate in this study and our study suggests that initiatives are needed to improve vaccine uptake in people with IMIDs. In patients with IBD, a similar pneumococcal vaccine uptake of 38% has been reported from a French online survey^10^ while a lower rate of 10.3% was reported from a gastro-enterology clinic in Canada^9^. Pneumococcal vaccine uptake in people with Axial SpA was reported to be 32.5% in Switzerland^17^. The multinational COMORA study reported higher uptake of pneumococcal vaccine in some countries e.g., France, but lower uptake in most countries^18^. These questionnaire studies are prone to bias from self-reported vaccination uptake.

Consistent with previous research on factors associated with vaccine uptake, female sex, increasing age and presence of an additional indication for vaccination significantly associated with receiving pneumococcal vaccination 11 19 20.

Barriers to vaccination have included the fear that vaccines may trigger IMID flare^12 21^. This study did not find a significant association between pneumococcal vaccination and flare of the underlying inflammatory disease. Similarly, a systematic review and meta-analysis of pneumococcal vaccine immunogenic studies in patients with SLE did not find an association between the vaccine and disease activity ^13^. Safety studies in the general population have shown that pneumococcal vaccine is well tolerated, particularly the pneumococcal polysaccharide vaccine (PPV23) which is recommended in people aged 65 years and older and individuals with underlying conditions including immunosuppression ^22^. Previous work by us reported no association between vaccination with the seasonal influenza vaccine and AIRD flare, and no association between vaccination against COVID-19 and AIRD, IBD, and psoriasis flare^23–26^. A meta-analysis of prior uncontrolled studies reported a 2% pooled prevalence of IBD flare with vaccination, however, it is not known if this was temporally related or coincidence^27^.

Strengths of this study include a large nationally representative sample of people with IMIDs in the UK given the almost universal registration with a GP for all UK residents^14^. We studied a wide range of IMIDs improving the generalizability of the findings. The use of primary-care prescription and consultation data minimised recall bias on the association between vaccination and disease flares. To improve the validity of our case definition, we used a combination of diagnostic and prescription codes to ascertain people with IMID. Additionally, we defined IBD flares according to a validated definition^15^ and we undertook a sensitivity analysis for the association between pneumococcal vaccination and AIRD flare using a flare definition validated for IBD flare. Furthermore, to improve outcome fidelity, we excluded participants with diagnoses that could potentially explain corticosteroid prescribed on the same date as the AIRD or IBD flare. Finally, our use of SCCS methodology controlled for between-person confounding which is a serious problem in observational studies of vaccine safety.

There are limitations to our study. Firstly, some vaccinations that were administered outside of primary-care for example in hospital or at the workplace as for health care professionals may not have been recorded in the CPRD, reducing vaccination uptake estimates. This is unlikely to have a significant impact on our results as vaccination is almost exclusively a general practice activity in the UK. Where non-primary care administration of the vaccine was recorded, it was excluded from the vaccine safety study as it is difficult to be sure of the date of vaccination in such instances. Second, the type of vaccine was not assessed as the vast majority of vaccinations were with the PPV23 vaccine, which has been universally used for risk groups since 1992^5^. Third, we were unable to assess the impact of biologics on vaccine safety because their prescription is not recorded in the CPRD. We see no reason though to expect more extreme immunologically driven side effects in these groups given the possibility of less immunogenic response with biologic use ^28^. Fourth, data on disease activity and flares managed in hospital or specialist clinics are not recorded in CPRD. This does not affect the validity of our findings because any resulting bias will be non-differential. Fifth, because our definition of AIRD and IBD flare required consultation and/or prescription, minor flares not needing drug treatment were not considered as an outcome in the vaccine safety study. It is possible that there may be an association of vaccination with minor flares that were not ascertained in our study. However, such effects would be unlikely to greatly discourage vaccination uptake and it is the more significant flares that we have studied which are of primary concern. Flares managed in hospital or specialist clinics were excluded.

In conclusion, this study provides recent UK-wide population evidence that the uptake of pneumococcal vaccine in people with IMIDs is suboptimal particularly in patients with IBD, those younger than 65 years in age, and in those without another indication for vaccination. It also demonstrated that pneumococcal vaccination does not associate with flare of the underlying IMIDs. These data should be used to promote pneumococcal vaccination in this at-risk population.

## Author contribution

The study was conceived by Prof Abhishek. All authors were involved in the design of the study. The analysis was carried out by Dr Nakafero. All authors edited the first and all subsequent drafts and approved the final draft for submission.

## Data sharing statement

Data used in the study are from the Clinical Practice Research Datalink (CPRD). Due to CPRD licencing rules, we are unable to share data used in this study with third parties. The data used in this study may be obtained directly from the CPRD. Study protocol is available from www.cprd.com.

## References

1. van Aalst M, Lötsch F, Spijker R, et al. Incidence of invasive pneumococcal disease in immunocompromised patients: A systematic review and meta-analysis. Travel Med Infect Dis 2018;24:89–100.

2. Wotton CJ, Goldacre MJ. Risk of invasive pneumococcal disease in people admitted to hospital with selected immune-mediated diseases: record linkage cohort analyses. J Epidemiol Community Health 2012;66(12):1177–81.

3. Shea KM, Edelsberg J, Weycker D, et al. Rates of pneumococcal disease in adults with chronic medical conditions. Open Forum Infect Dis 2014;1(1):ofu024.

4. Shigayeva A, Rudnick W, Green K, et al. Invasive Pneumococcal Disease Among Immunocompromised Persons: Implications for Vaccination Programs. Clin Infect Dis 2016;62(2):139–47.

5. UK Health Security Agency. Pneumococcal: the green book, chapter 25, 2023.

6. Lopez A, Mariette X, Bachelez H, et al. Vaccination recommendations for the adult immunosuppressed patient: A systematic review and comprehensive field synopsis. Journal of Autoimmunity 2017;80:10–27.

7. Campling J, Vyse A, Liu H-H, et al. A review of evidence for pneumococcal vaccination in adults at increased risk of pneumococcal disease: risk group definitions and optimization of vaccination coverage in the United Kingdom. Expert Review of Vaccines 2023;22(1):785–800.

8. Costello R, Winthrop KL, Pye SR, et al. Influenza and Pneumococcal Vaccination Uptake in Patients with Rheumatoid Arthritis Treated with Immunosuppressive Therapy in the UK: A Retrospective Cohort Study Using Data from the Clinical Practice Research Datalink. PLoS One 2016;11(4):e0153848.

9. Malhi G, Rumman A, Thanabalan R, et al. Vaccination in inflammatory bowel disease patients: attitudes, knowledge, and uptake. J Crohns Colitis 2015;9(6):439–44.

10. Loubet P, Verger P, Abitbol V, et al. Pneumococcal and influenza vaccine uptake in adults with inflammatory bowel disease in France: Results from a web-based study. Dig Liver Dis 2018;50(6):563–67.

11. Loubet P, Kernéis S, Groh M, et al. Attitude, knowledge and factors associated with influenza and pneumococcal vaccine uptake in a large cohort of patients with secondary immune deficiency. Vaccine 2015;33(31):3703–08.

12. Fuller A, Hancox J, Vedhara K, et al. Barriers and facilitators to vaccination uptake against COVID-19, influenza, and pneumococcal pneumonia in immunosuppressed adults with immune-mediated inflammatory diseases: A qualitative interview study during the COVID-19 pandemic. PLoS One 2022;17(9):e0267769.

13. Pugès M, Biscay P, Barnetche T, et al. Immunogenicity and impact on disease activity of influenza and pneumococcal vaccines in systemic lupus erythematosus: a systematic literature review and meta-analysis. Rheumatology (Oxford) 2016;55(9):1664–72.

14. Herrett E, Gallagher AM, Bhaskaran K, et al. Data Resource Profile: Clinical Practice Research Datalink (CPRD). International journal of epidemiology 2015;44(3):827–36.

15. Lewis JD, Aberra FN, Lichtenstein GR, et al. Seasonal variation in flares of inflammatory bowel disease. Gastroenterology 2004;126(3):665–73.

16. Nakafero G, Grainge MJ, Myles PR, et al. Association between inactivated influenza vaccine and primary care consultations for autoimmune rheumatic disease flares: a self-controlled case series study using data from the Clinical Practice Research Datalink. Annals of the Rheumatic Diseases 2019;78(8):1122–26.

17. Stoffel ST, Colaninno A, Bräm R, et al. Pneumococcal vaccination among adult risk patient with axial spondyloarthritis in Switzerland: Data from the survey of the ankylosing spondylitis association of Switzerland (SVMB). Vaccine 2022;40(43):6206–10.

18. Hmamouchi I, Winthrop K, Launay O, et al. Low rate of influenza and pneumococcal vaccine coverage in rheumatoid arthritis: data from the international COMORA cohort. Vaccine 2015;33(12):1446–52.

19. Nakafero G, Grainge MJ, Myles PR, et al. Predictors and temporal trend of flu vaccination in auto-immune rheumatic diseases in the UK: a nationwide prospective cohort study. Rheumatology (Oxford) 2018;57(10):1726–34.

20. Garg M, Mufti N, Palmore TN, et al. Recommendations and barriers to vaccination in systemic lupus erythematosus. Autoimmunity reviews 2018;17(10):990–1001.

21. Lawson EF, Trupin L, Yelin EH, et al. Reasons for failure to receive pneumococcal and influenza vaccinations among immunosuppressed patients with systemic lupus erythematosus. Semin Arthritis Rheum 2015;44(6):666–71.

22. Musher DM, Manof SB, Liss C, et al. Safety and antibody response, including antibody persistence for 5 years, after primary vaccination or revaccination with pneumococcal polysaccharide vaccine in middle-aged and older adults. J Infect Dis 2010;201(4):516–24.

23. Adams L, Nakafero G, Grainge MJ, et al. Is vaccination against COVID-19 associated with psoriasis or eczema flare? Self-controlled case series analysis using data from the Clinical Practice Research Datalink (Aurum). Br J Dermatol 2023;188(2):297–99.

24. Card TR, Nakafero G, Grainge MJ, et al. Is Vaccination Against COVID-19 Associated With Inflammatory Bowel Disease Flare? Self-Controlled Case Series Analysis Using the UK CPRD. Am J Gastroenterol 2023;118(8):1388–94.

25. Nakafero G, Grainge MJ, Card T, et al. Is vaccination against COVID-19 associated with autoimmune rheumatic disease flare? A self-controlled case series analysis. Rheumatology (Oxford) 2023;62(4):1445–50.

26. Nakafero G, Grainge MJ, Myles PR, et al. Association between inactivated influenza vaccine and primary care consultations for autoimmune rheumatic disease flares: a self-controlled case series study using data from the Clinical Practice Research Datalink. Ann Rheum Dis 2019;78(8):1122–26.

27. Desalermos A, Pimienta M, Kalligeros M, et al. Safety of Immunizations for the Adult Patient With Inflammatory Bowel Disease-A Systematic Review and Meta-analysis. Inflamm Bowel Dis 2022;28(9):1430–42.

28. UK Health Security Agency. Immunisation of individuals with underlying medical conditions, 2023.

