## supplementary material for "Uptake and safety of pneumococcal vaccination in adults with immune mediated inflammatory diseases: a nationwide observational study using data from the Clinical Practice Research Datalink (Gold) in the UK"

Table S1: Code list Pneumococcal vaccination and vaccines

**Medcodes**

| medcode | readcode | readterm |
|---|---|---|
| 5764 | 7L19700 | Subcutaneous injection of Pneumovax II |
| 11363 | 6572 | Pneumococcal vaccination |
| 30411 | 9Oo..00 | Pneumococcal vaccination administration |
| 36826 | 6572000 | Pneumococcal vaccination given |
| 53198 | 657K.00 | Booster pneumococcal vaccination |
| 97759 | 657P.00 | Pneumococcal vaccination given by other healthcare provider |

**Product codes**

| prodcode | Description |
|---|---|
| 821 | Pneumococcal polysaccharide conjugated vaccine (adsorbed) suspension for injection 0.5ml vials |
| 832 | Pneumococcal polysaccharide vaccine solution for injection 0.5ml vials |
| 930 | Pneumovax II vaccine solution for injection 0.5ml vials (sanofi pasteur MSD Ltd) |
| 1327 | PNEUMOVAX VAC |
| 3684 | Pnu-Imune vaccine solution for injection 0.5ml vials (Wyeth Pharmaceuticals) |
| 15482 | PNEUMOVAC PLUS VACCINE VAC |
| 42602 | Prevenar 13 vaccine suspension for injection 0.5ml pre-filled syringes (Pfizer Ltd) |
| 42612 | Pneumococcal polysaccharide conjugated vaccine (adsorbed) suspension for injection 0.5ml pre-filled syringes |
| 42991 | Pneumococcal 10-valent saccharide conjugated absorbed vaccine |
| 50264 | Pneumovax II vaccine solution for injection 0.5ml pre-filled syringes (sanofi pasteur MSD Ltd) |
| 50338 | Prevenar vaccine suspension for injection 0.5ml pre-filled syringes (Pfizer Ltd) |
| 53159 | Synflorix vaccine suspension for injection 0.5ml pre-filled syringes (GlaxoSmithKline UK Ltd) |
| 4530007 | PNEUMOVAX VAC |
| 6007001 | PNEUMOCOCCAL 23-VALENT POLYSACCHARIDE vaccine |
| 6008001 | PNEUMOVAX II vaccine |
| 7920001 | PNEUMOCOCCAL 7-VALENT SACCHARIDE CONJUGATED adsorbed vaccine |
| 9289001 | PREVENAR vaccine |
| 10904001 | PNU-IMUNE vaccine |

Table S2: Code list joint pain codes.

| Medcode | Readcode | Readterm |
|---|---|---|
| 5899 | 1A53.12 | C/O - lumbar pain |
| 29402 | N094411 | Hand joint pain |
| 1330 | N094512 | Hip joint pain |
| 9105 | 1D13100 | C/O - pain in hallux |
| 1274 | N33A100 | Clavicle pain |
| 374 | 182..00 | Chest pain |
| 11544 | N242300 | Neuropathic pain |
| 6166 | N094W00 | Anterior knee pain |
| 11962 | 1M00.00 | Pain in elbow |
| 1339 | N245011 | Thumb pain |
| 5476 | N12..13 | Acute back pain – disc |
| 4627 | N245100 | Foot pain |
| 4706 | N33A000 | Bony pelvic pain |
| 19020 | 25C..13 | O/E - lumbar pain on palpation |
| 1646 | N094211 | Elbow joint pain |
| 123 | N131.00 | Cervicalgia - pain in neck |
| 54791 | N094D11 | Elbow joint pain |
| 10231 | 16C9.00 | Chronic low back pain |
| 1335 | N142.00 | Pain in lumbar spine |
| 14886 | N094711 | Ankle joint pain |
| 2781 | 1A59.00 | C/O pelvic pain |
| 6520 | N245000 | Hand pain |
| 5916 | N141.11 | Acute back pain – thoracic |
| 6313 | 1D13111 | C/O - pain in big toe |
| 9698 | 1824 | Anterior chest wall pain |
| 35744 | N131.11 | Pain in cervical spine |
| 8309 | N096.12 | Musculoskeletal pain – joints |
| 4544 | N245.00 | Pain in limb |
| 5787 | N245700 | Shoulder pain |
| 3322 | N245012 | Finger pain |
| 4948 | N141.00 | Pain in thoracic spine |
| 1286 | R040z11 | [D]Jaw pain |
| 9811 | R090G11 | [D] Pelvic pain |
| 917 | N33A.00 | Bone pain |
| 2494 | N245.15 | Heel pain |
| 2025 | N245.14 | Hand pain |
| 198 | N245.17 | Shoulder pain |
| 1219 | N245.16 | Leg pain |
| 855 | N245.13 | Foot pain |
| 822 | N245.12 | Arm pain |
| 1866 | N245.18 | Thigh pain |
| 2521 | N094311 | Wrist joint pain |
| 1258 | N245.11 | Ankle pain |
| 36132 | 2H45.00 | O/E - joint movement painful |
| 6644 | 1D13000 | C/O - pain in toes |
| 10389 | 1M12.00 | Anterior knee pain |
| 9517 | 1M10.00 | Knee pain |
| 554 | N094611 | Knee joint pain |
| 5762 | N245300 | Pain in arm |
| 7294 | N240300 | Rheumatic pain |
| 108855 | 1M02.00 | Shoulder joint painful on movement |
| 99477 | 1M01.00 | Pain in wrist |

| | | |
|---|---|---|
| 109347 | 1M03.00 | Shoulder joint painful on external rotation |
| 19687 | 1M0..00 | Pain in upper limb |
| 11979 | 1M00.11 | Elbow pain |
| 1946 | N245111 | Toe pain |
| 96084 | N094F11 | Wrist pain |
| 2397 | N094111 | Shoulder joint pain |
| 11609 | 1M1..00 | Pain in lower limb |
| 286 | N094K12 | Hip pain |
| 19322 | 1M13.00 | Ankle pain |
| 17799 | 1M11.00 | Foot pain |
| 5780 | N245200 | Pain in leg |

Table S3: Alternative indication for corticosteroid prescription

A: Codes used to define alternative indication for prescribing corticosteroids in the IBD flare and pneumococcal vaccinated patient cohort.

| Alternative indication ID | Number of codes per indication | Medcode | Readcode | Readterm |
|---|---|---|---|---|
| 1 | 1 | 709 | C34..00 | Gout |
| 2 | 1 | 1408 | N20..00 | Polymyalgia rheumatica |
| 3 | 1 | 292 | 1719.00 | Chesty cough |
| 4 | 1 | 92 | 171..00 | Cough |
| 5 | 1 | 312 | H060.00 | Acute bronchitis |
| 6 | 1 | 5861 | 2326.00 | O/E - expiratory wheeze |
| 7 | 2 | 5767 | 1B1G.11 | C/O - a headache |
| 8 | | 1958 | 8H53.00 | ENT referral |
| 9 | 4 | 538 | 9877.00 | Minor surgery done - injection |
| 10 | | 3546 | N215700 | Trochanteric bursitis |
| 11 | | 209 | 7K6Z200 | Injection of therapeutic substance into joint |
| 12 | | 1013 | N223.00 | Bursitis NOS |
| 13 | 5 | 23663 | 17ZZ.00 | Respiratory symptom NOS |
| 14 | | 7191 | 663P.00 | Asthma limiting activities |
| 15 | | 10043 | 66YJ.00 | Asthma annual review |
| 16 | | 4442 | H33z.00 | Asthma unspecified |
| 17 | | 7416 | 663N.00 | Asthma disturbing sleep |
| 18 | 8 | 13173 | 663O.00 | Asthma not disturbing sleep |
| 19 | | 10274 | 8B3j.00 | Asthma medication review |
| 20 | | 81 | 663..11 | Asthma monitoring |
| 21 | | 81 | 663..11 | Asthma monitoring |
| 22 | | 13174 | 663Q.00 | Asthma not limiting activities |
| 23 | | 42824 | 663q.00 | Asthma daytime symptoms |
| 24 | | 1649 | 9....00 | Administration |
| 25 | | 10274 | 8B3j.00 | Asthma medication review |

B: Codes used to define alternative indication for prescribing corticosteroids in the AIRD flare and pneumococcal vaccinated patient cohort.

| Alternative indication ID | Number of codes per indication | Medcode | Readcode | Readterm (Alternative indication) |
|---|---|---|---|---|
| 1 | 1 | 5175 | 173..11 | Breathlessness symptom |
| 2 | 1 | 292 | 1719 | Chesty cough |
| 3 | 1 | 2891 | 1737.11 | Wheezing symptom |
| 4 | 1 | 1272 | M28..00 | Urticaria |
| 5 | 1 | 312 | H060.00 | Acute bronchitis |
| 6 | 1 | 2581 | H06z000 | Chest infection NOS |
| 7 | 1 | 1001 | H3...00 | Chronic obstructive pulmonary disease |
| 8 | 1 | 78 | H33..00 | Asthma |
| 9 | 1 | 185 | H333.00 | Acute exacerbation of asthma |
| 10 | 1 | 1784 | J41..12 | Ulcerative colitis and/or proctitis |
| 11 | 1 | 5349 | 173..13 | Shortness of breath symptom |
| 12 | 1 | 293 | H06z111 | Respiratory tract infection |
| 13 | 1 | 173 | 1737 | Wheezing |
| 14 | 1 | 7884 | H3y1.00 | Chron obstruct pulmonary dis wth acute exacerbation, unspec |
| 15 | 1 | 68 | H06z011 | Chest infection |
| 16 | 4 | 11287 | 66YM.00 | Chronic obstructive pulmonary disease annual review |
| 17 | | 19427 | 173I.00 | MRC Breathlessness Scale: grade 2 |
| 18 | | 101042 | 8BMW.00 | Issue of chronic obstructive pulmonary disease rescue pack |
| 19 | | 10802 | H37..00 | Moderate chronic obstructive pulmonary disease |
| 20 | 6 | 28743 | 66Yf.00 | Number of COPD exacerbations in past year |
| 21 | | 19427 | 173I.00 | MRC Breathlessness Scale: grade 2 |
| 22 | | 10337 | 68M..00 | Spirometry screening |
| 23 | | 11287 | 66YM.00 | Chronic obstructive pulmonary disease annual review |
| 24 | | 8484 | 663H.00 | Inhaler technique - good |
| 25 | | 13683 | 8HRC.00 | Referral for spirometry |
